# Asymptotic solutions of the SIR and SEIR models well above the epidemic threshold

**DOI:** 10.1101/2021.03.24.21254226

**Authors:** Gregory Kozyreff

## Abstract

A simple and explicit expression of the solution of the SIR epidemiological model of Kermack and McKendrick is constructed in the asymptotic limit of large basic reproduction numbers R_0_. The proposed formula yields good qualitative agreement already when R_0_ ⩾ 3 and rapidly becomes quantitatively accurate as larger values of R_0_ are assumed. The derivation is based on the method of matched asymptotic expansions, which exploits the fact that the exponential growing phase and the eventual recession of the outbreak occur on distinct time scales. From the newly derived solution, an analytical estimate of the time separating the first inflexion point of the epidemic curve from the peak of infections is given. Finally, we use the same method on the SEIR model and find that the inclusion of the “exposed” population can dramatically alter the time scales of the outbreak.

## 1. Introduction

The COVID-19 pandemic has impacted all aspects of our daily lives. It has thus triggered an extraordinary world-wide response across all fields of science, from virology and pharmacology to sociology. Within the scientific crowd, applied mathematicians and theoretical physicists also brought their contribution, notably by using their mathematical expertise in epidemiological modelling Bagal et al. (2020); Roda et al. (2020); Calafiore et al. (2020); Simha et al. (2020); Casares & Khan (2020); Chen et al. (2020); Chowdhury et al. (2020); Gatto et al. (2020); Bastos & Cajueiro (2020); Giordano et al. (2020); Oliveira et al. (2021); Merbis (2021). Deterministic compartment models allow one in principle to develop an accurate global view of the contagion dynamics within large populations. Compartments can be used to divide the population into age categories as well as according to the evolution of the illness, for those who have been infected Brauer et al. (2012); Gatto et al. (2020); Giordano et al. (2020). These multiple levels of description are useful and necessary, and can be made to closely reproduce the data. However, they contain a plethora of fitting parameters and can therefore be complex to interpret. Fortunately, it turns out that the simplest of all compartment models, SIR, faithfully reproduces the global dynamics at the level of a country or a large city with COVID-19 Bastos & Cajueiro (2020); Kozyreff (2021). As a result, the SIR model remains a useful tool at such global level of description. Besides, epidemiologic thinking can be used, either with the SIR model or variants thereof, to model the spread of rumours Daley & Kendall (1964), songs Rosati et al. (2021) or singer popularity Tweedle & Smith (2012) as well as fashion Acerbi et al. (2012) and fads on the internet Bauckhage et al. (2013), making the SIR model an active subject of research, nearly a century after having been put forward by Kermack & McKendrick (1927). We write it as follows

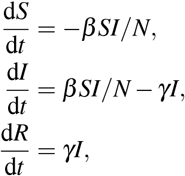

where *S, I*, and *R* respectively denote the number of susceptible, infected and removed individuals, with constant sum *N* = *S* + *I* + *R*. The population *R* includes both those who have recovered from the illness and those who have died. Above, *β* is the contact rate, *γ* is the recovery rate, and their ratio

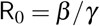

is the basic reproduction number. Despite its long history, there continues to be many efforts to try and solve it analytically Brauer (2005); Khan et al. (2009); Harko et al. (2014); Bohner et al. (2019); Barlow & Weinstein (2020); Giubilei (2020); Kröger & Schlickeiser (2020); Mieghem (2020); Prodanov (2020); de Souza (2021); Kudryashov et al. (2021). In their seminal paper, Kermack and McKendrick offered an approximate solution in the limit of a “small epidemic”, *i*.*e*. when the reproduction number R_0_ is just above 1. So far, this has remained the only explicit approximate analytical solution available, in the sense that it can be justified as an asymptotic limit of the true solution. Well above the epidemic threshold, Kendall showed in 1956 that the solution can be written in parametric form Kendall (1956). Unfortunately, it is only given implicitly, such that time is expressed in terms of one of the dynamical variables through an integral [see Eq. (2.9) below.] Hence, this solution is not directly interpretable and its analysis can be quite involved Mieghem (2020). Alternatives have been proposed in the form of converging series Khan et al. (2009); Barlow & Weinstein (2020), but these can involve expansions with as many as 15 up to 60 terms and, hence, are again impractical for analysis. More down-to-earth is the approach by which the solution is fitted by a well-chosen ansatz with a few parameters Clark et al. (2020); de Souza (2021). However, these ansatzes are not derived from the model and their parameters must be fitted to the data, rather than deduced from the model. During the first wave of the COVID-19 pandemic, and before confinement measures were imposed, values of R_0_ between 3 and more than 6 have been estimated Nadeau et al. (2021); Liu et al. (2020) and recovery times on the order of two weeks for patients admitted in hospital have been estimated Kozyreff (2021), suggesting *γ* = 1*/*15 day^−1^ (for the first wave of the pandemic). In other context, much larger values of R_0_ have been inferred Rosati et al. (2021).

### 1.1 Statement of results

The aim of this paper is to show that

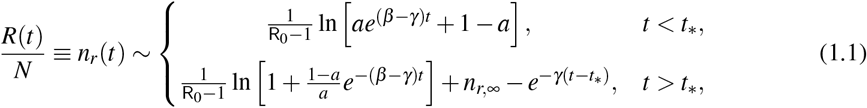

where *a* is the infected fraction of the population at *t* = 0, while *n*_*r*,∞_ and *t*_∗_ are given by

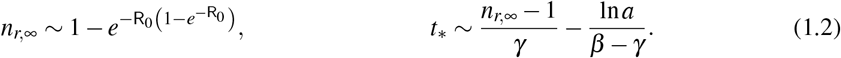

Given *n*_*r*_(*t*), one may infer the infected fraction of the population exactly as

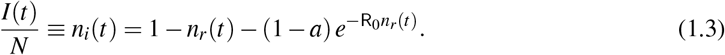

Eq. (1.1) is remarkably simple and compact. The “∼ “ sign in it indicates that it holds asymptotically in the limit R_0_ »1. However, a good qualitative agreement with the numerical solution is already found for R_0_ = 3 and the approximate formula rapidly becomes as good as exact with increasing R_0_, see Fig. 1. Hence, Eq. (1.1) complements the classical “small epidemic” formula derived by Kermack & McKendrick (1927), which is valid in the limit 0 *<* R_0_ −1 «1. When, in 1956, D. G. Kendall presented the implicit analytical solution [Eq. (2.9) below] of the SIR model, he noted:

**FIG. 1.**
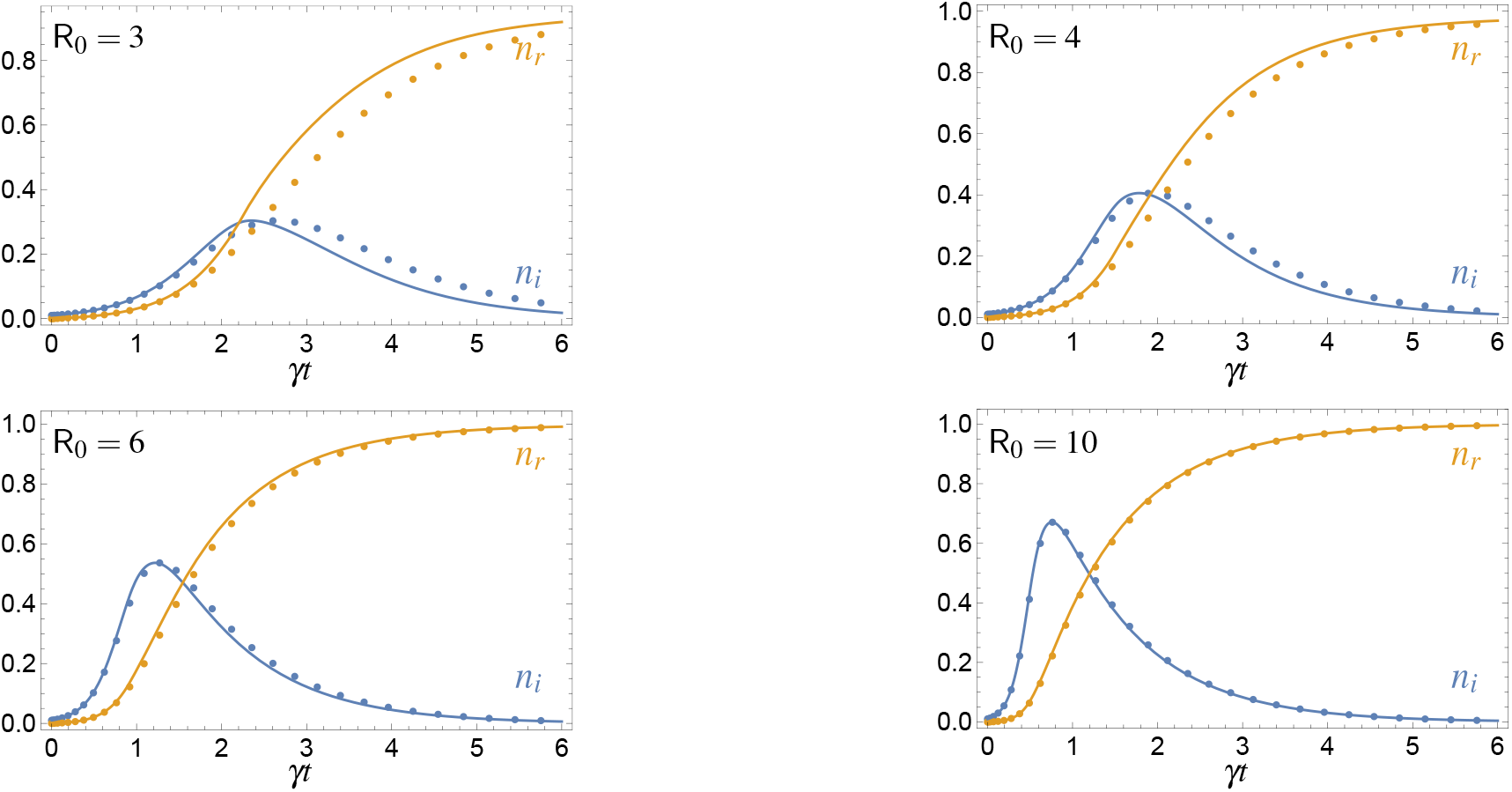
Comparison between the approximate formulas (1.1) and (1.3) (full lines) and the numerical solution of the SIR equation (dotted lines) for several values of the reproduction number R_0_. *n*_*i*_(0) = *a* = 0.01. Note that, once R_0_ is given, the parameter *γ* only appears through the dimensionless product *γt*.

“*It is curious that the Kermack and McKendrick approximation should have been accepted without comment for nearly thirty years; the exact solution is easily obtained and the difference between the two can be of practical significance*.” (Kendall, 1956, p. 150)

Similarly, the reader will find that the derivation of the present asymptotic formula is quite simple and could have been established much earlier with well-established asymptotic techniques.

As an illustration of the usefulness of the present analytical expression, let us compute the time elapsed between two important moments of the epidemic, namely the time *t*_1_ of first inflexion of the curve *n*_*i*_(*t*) and the time *t*_2_ of the peak of the epidemic. In appendix, we show that the former happens when *n*_*r*_ is equal to *n*_1_, solution of

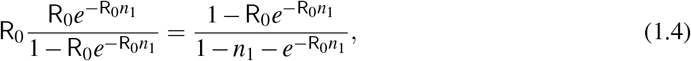

while the latter happens when

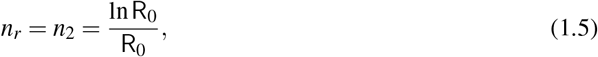

in the limit *a* → 0. It turns our that *t*_1_ *< t*_∗_ and that *t*_2_ *> t*_∗_. Hence, using Eq. (1.1), we have

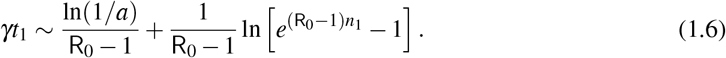

Meanwhile, for *t*_2_, we have

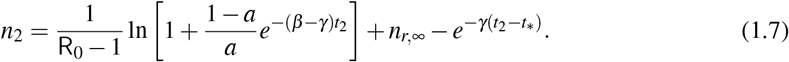

Above, we may assume in first approximation that *t*_2_ is sufficiently large that the logarithmic term is negligible. Thus, we obtain the more manageable expression

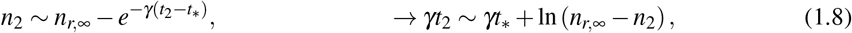

and find

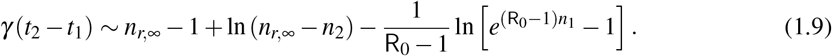

A comparison between this approximate formula and the numerics is given in Fig. 2, suggesting that the relative error associated to Eq. (1.9) is *O* [(R_0_ ln R_0_)^−1^]. Collecting data in real-time allows one to detect when the slope of the epidemic curve has reached a maximum; *t*_2_ − *t*_1_ is then the time left before the epidemic starts to recede. In a context such as the first wave of the COVID-19 pandemic, being able to predict when the epidemic will reach its peak can be a valuable asset both to manage health resources and to communicate to the public.

**FIG. 2.**
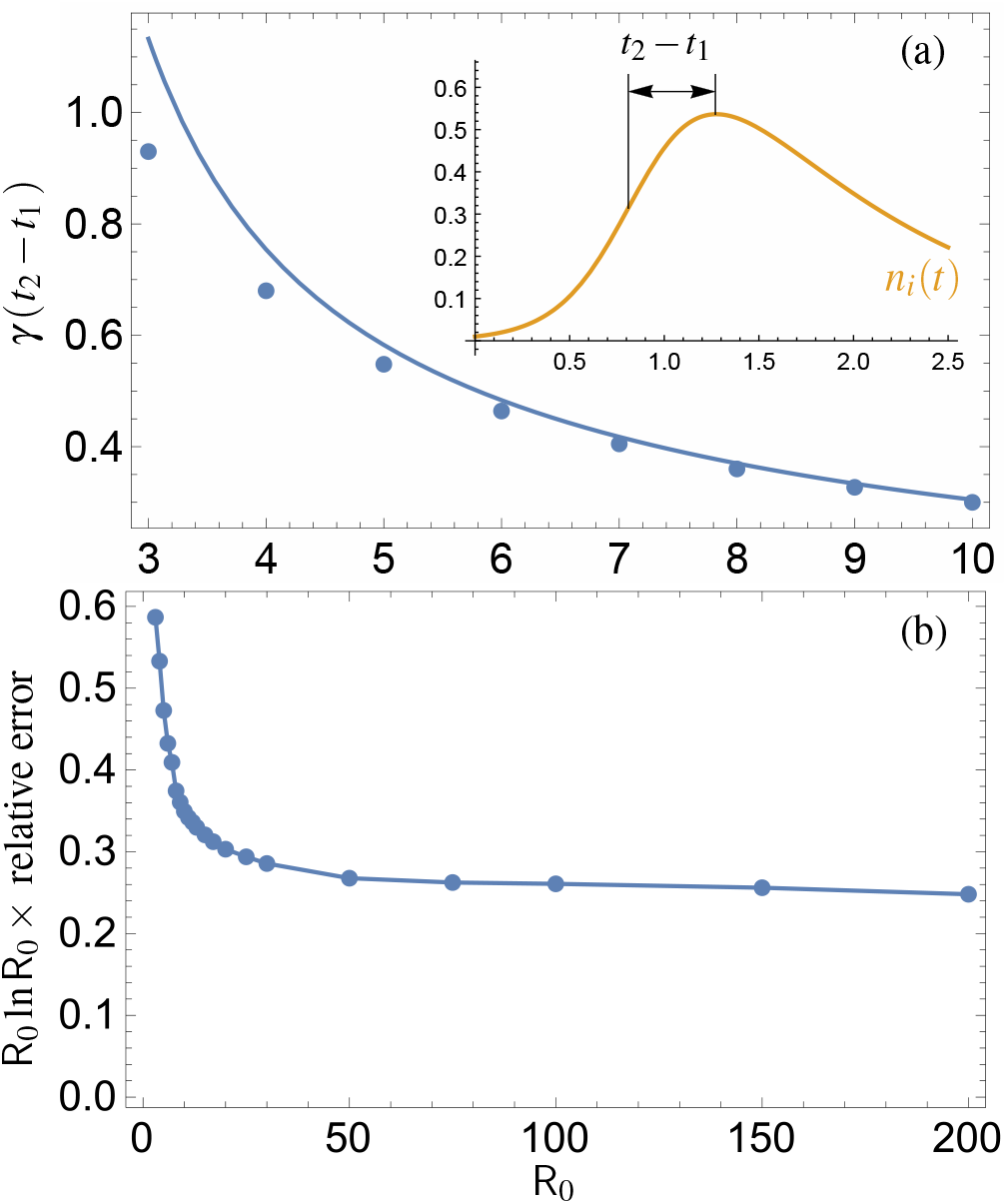
Time from inflexion to epidemic maximum as a function of R_0_. (a) Comparison between numerical solution (dots) and asymptotic formula (full line). (b) Plot of the relative error |*y*_num_*/y*_asy_ − 1| multiplied by R_0_ ln R_0_, where *y*_num_ and *y*_asym_ designate the numerical value and asymptotic approximation of *t*_2_ − *t*_1_, respectively.

## 2. Preliminaries

Before embarking into the asymptotic analysis of the SIR model, we first recall a few well-known facts. Eliminating *S* thanks to the relation *S*(*t*) + *I*(*t*) + *R*(*t*) = *N*, the evolution equations for the fractions *n*_*i*_ = *I/N* and *n*_*r*_ = *R/N* are

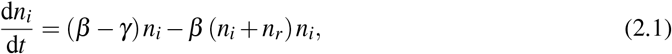

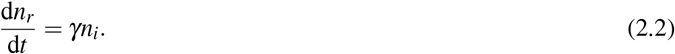

They should be solved with the initial condition

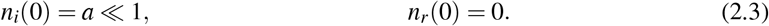

At the start of the outbreak, *n*_*i*_, *n*_*r*_ « 1 and the linearised evolution equations immediately yield

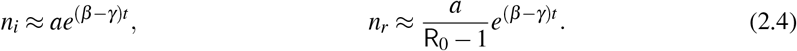

(Here, the ‘≈’ sign is used because nonlinear terms in Eqs. (2.1) and (2.2) are neglected.) On the other hand, dividing Eq. (2.1) by Eq. (2.2), one obtains

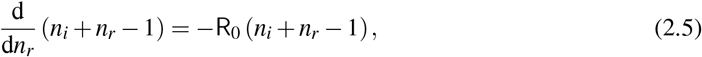

which directly yields Eq. (1.3). Thanks to the explicit dependence *n*_*i*_[*n*_*r*_(*t*)], Eq. (2.2) eventually becomes a first-order nonlinear ODE:

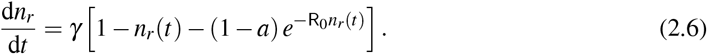

As *t* → ∞, *n*_*r*_ tends to the value *n*_*r*,∞_ that makes the right hand side vanish:

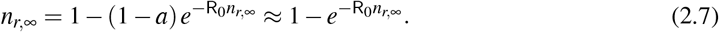

In the large-R_0_ limit, the second term in the right hand side above is small, so that a recursive resolution can be set up. Note that, since *a* «1, as is generically the case, we neglect it in (2.7). An excellent approximation is given by

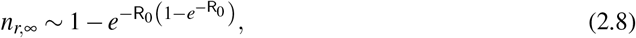

with an absolute error of less than 0.002 for R_0_ *>* 3. Alternatively, one may express *n*_*r*,∞_ in terms of the tabulated Lambert function Mieghem (2020).

It is immediate to see that Eq. (2.6) can formally be integrated as

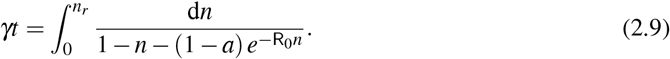

This expression has been known and studied for a long time Kendall (1956). Note that if both *γ* and *β* are functions of time, such that their ratio R_0_ = *β/γ* remains constant, then Eq. (2.9) remains valid, with *γt* replaced by the effective time *τ* =∫ *γ*(*t*)d*t* Kröger & Schlickeiser (2020). Recently, a thorough analysis of the integrand of Eq. (2.9) was carried out, yielding upper and lower bounds for the function *t*(*n*_*r*_) Mieghem (2020). In addition, series representations of the integral based either on Taylor expansion of the integrand near the origin or based on its poles in the complex-*n* plane were given in that work. The first of these approaches effectively amounts to assume that *n* is small and thus generalizes Kermack and McKendrick approximate solution. However, the formulas in Mieghem (2020) are rather involved, making the inversion of the relation *t*(*n*_*r*_) laborious.

## 3. Matched asymptotic expansions

The method consists in deriving separate approximations at the early stage and at the later stage of the outbreak. One the one hand, the outer solution has a characteristic time *γ*^−1^ and describes the decaying phase of the epidemic. On the other hand, the inner solution applies to the growing phase and evolves on a shorter time scale. In the language of asymptotics, the R_0_ »1 limit produces a *boundary layer* problem Bender & Orszag (1999). The two approximations are constrained by matching conditions at intermediate stages of the outbreak. Thanks to the fact that they have an overlapping range of validity, one can then construct a composite solution, Eq. (1.1), that is uniformly valid in time.

### 3.1 Outer solution

When 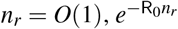 is exponentially small and can at first be neglected in Eq. (2.6). We thus have

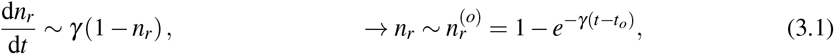

where the superscript (*o*) refers to “outer” approximation and *t*_*o*_ is an arbitrary constant. However, knowing the actual limiting value of *n*_*r*_ as *t* → ∞, a better approximation is

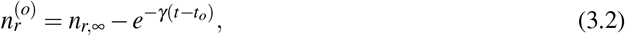

where *n*_*r*,∞_ is given in Eq. (2.8). Since *n*_*r*,∞_ and 1 only differ by an exponentially small quantity, they are asymptotically equivalent as R_0_ → ∞. However, the correction brought about by using *n*_*r*,∞_ in Eq. (3.2) considerably improves the approximation for moderately large R_0_ and thus expands the range of usability of the final result, Eq. (1.1).

Note that 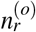 is only defined for *t > t*, since *n*_r_ must be a positive number.

### 3.2 Inner solution

At earlier stages of the epidemics, *n*_*r*_ is *O* (R_0_^−1^). More precisely, Eq. (2.4) suggests to write

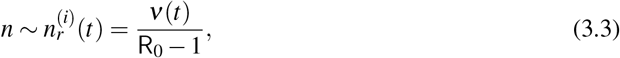

where (*i*) refers to “inner” approximation. Then Eq. (2.6) becomes

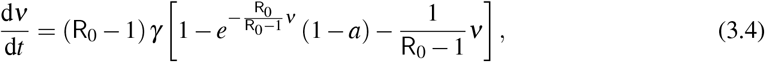

Unlike Eq. (2.6), this last equation can be integrated explicitly:

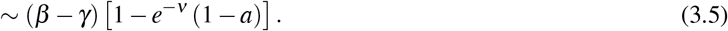

where we used the fact that *ν*(0) = 0. Hence,

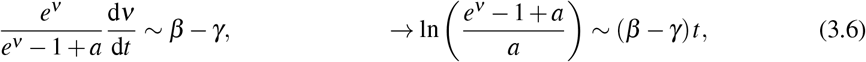

and

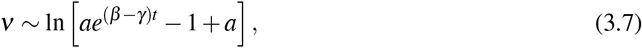

Just as we used *n*_*r*,∞_ instead of 1 in the outer solution, we here and henceforth use 1*/*(R_0_ −1) instead of 1*/*R_0_: the two are asymptotically equivalent to leading order in R_0_, but the former yields superior numerical approximation, on account of the linear expression (2.4).

### 3.3 Matching

So far, we have derived two approximations that efficiently describe the solution at different moments of the epidemic. However, the constant *t*_*o*_ in Eq. (3.2) is still unknown. To determine it, we must impose that (3.2) and (3.8) match in an intermediate region. Loosely speaking, one wants that

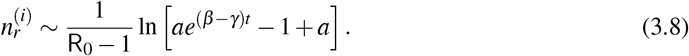

To formalise this statement more precisely, let us define the matching region as

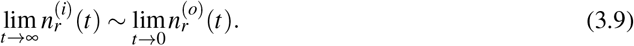

where *η* is such that *γηΔt* « 1, while (*β* − *γ*) *ηΔt* » 1. This double asymptotic constraint is possible since (*β* − *γ*) */γ* = R_0_ − 1 » 1 by assumption. On the one hand, we have

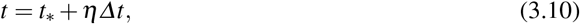

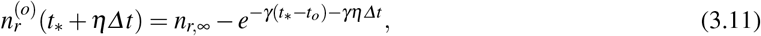

On the other hand, the inner solution yields

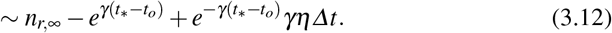

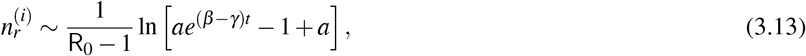

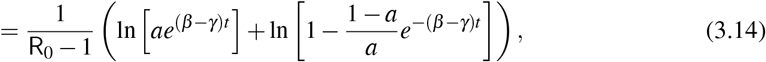

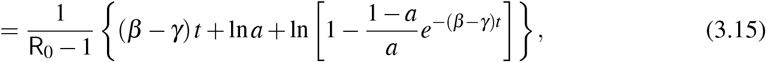

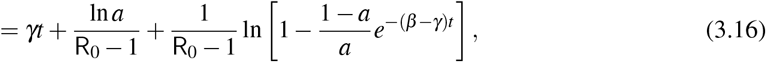

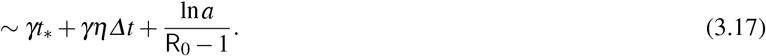

Equating expressions (3.12) and (3.17), we obtain the two conditions

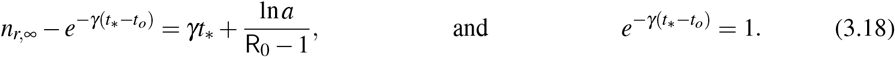

Hence, we have *t*_*o*_ = *t*_∗_ and

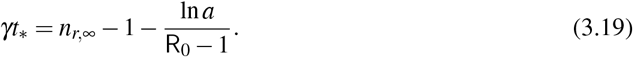

### 3.4 Composite solution

As indicated above, 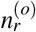 is only defined for *t > t* = *t*. For *t < t*, only the inner solution holds:

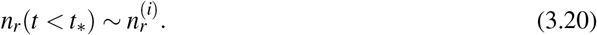

On the other hand, for *t > t*_∗_, we may construct a uniformly valid approximation as

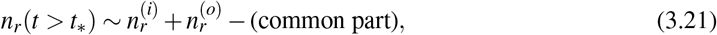

where the common part is the time-dependance that 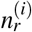 and 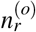 have in common in the matching region, and which should be removed to avoid double counting. One has, from (3.17),

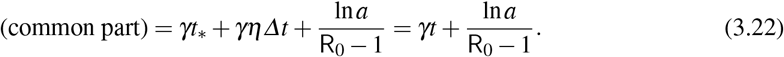

Hence

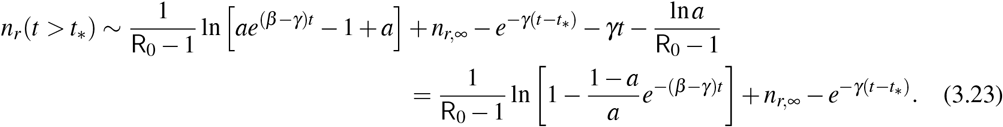

Eqs. (3.20) and (3.23) yield Eq. (1.1), which completes the derivation.

## 4. Extension to the SEIR model

The method of matched asymptotic expansions can be applied to more elaborate models as well. We demonstrate this on the SEIR model, which extends the SIR by adding a population of contaminated individuals that are not contagious yet. This population is called “exposed” and designated by *E*. Once a person is exposed to the disease, it takes an average time 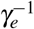 before he or she falls into the category *I* of infected people. The SEIR model reads as follows

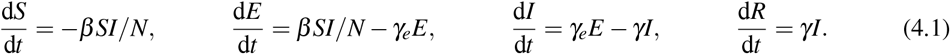

As before, *S*(*t*) + *E*(*t*) + *I*(*t*) + *R*(*t*) = *N*, allowing us to eliminate *S*(*t*). Defining the fractions *n*_*e*_ = *E/N, n*_*i*_ = *I/N* and *n*_*r*_ = *R/N*, we thus obtain

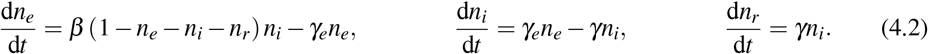

We propose to solve it with the initial conditions *n*_*i*_(0) = *a*« 1, *n*_*e*_(0) = *n*_*r*_(0) = 0. The introduction of the new variable *n*_*e*_ comes with a new parameter *γ*_*e*_. We discard the situation *γ*_*e*_» *β, γ*, which corresponds to the limit of a vanishing duration of the “exposed” state. In that limit, it is easy to find that *γ*_*e*_*n*_*e*_ *∼ β* (1− *n*_*i*_ −*n*_*r*_), which leads us back to the SIR model. Rather, we focus on the limit *β* » *γ*_*e*_, *γ* and assume that *γ*_*e*_ is larger than, but of the same order of magnitude as, *γ*. Following exactly the same reasoning as in Sec. 2, one easily derives the first integral

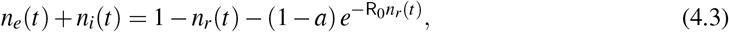

where R_0_ » 1 is still defined as the ratio *β/γ*.

### 4.1 Outer region

In the outer region, *n*_r_ = *O*(1) and Eq. (4.3) yields 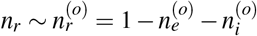. We are thus left with

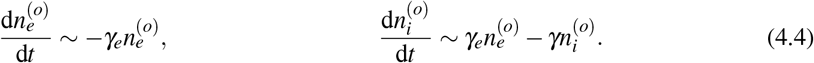

The solution is

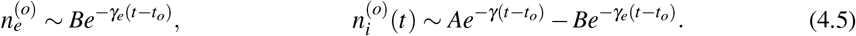

where *t*_*o*_, *A*, and *B* are constants to be determined by matching.

### 4.2 Inner region

In the inner region, we find the appropriate units and time scale by writing

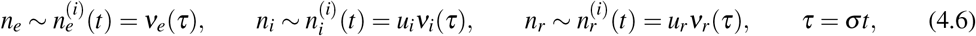

where the constants *u*_*i*_, *u*_*r*_ and *σ* are arbitrary. Eqs. (4.2) become

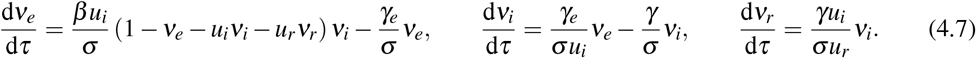

Let us choose *σ, u*_*i*_ and *u*_*r*_ such that *βu*_*i*_*/σ* = 1, *γ*_*e*_*/*(*σu*_*i*_) = 1 and *γu*_*i*_*/*(*σu*_*r*_) = 1, that is

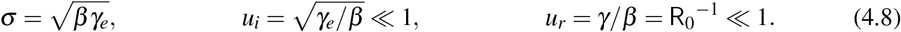

Hence, to leading order, the first two of Eqs. (4.7) reduce to

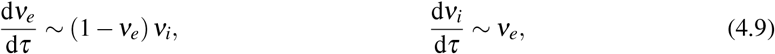

where we neglected 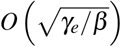and *O* (R_0_^−1^)terms. Equivalently, we have

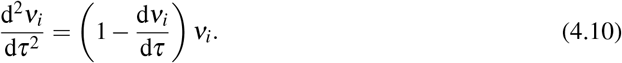

The above equation should be solved with the initial conditions

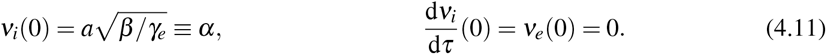

There is only one parameter left in this problem: *α*. Let us denote the numerical solution of this problem as

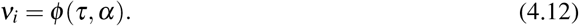

It does not seem possible to express this function in closed form. However, since it depends only on *τ* and *α*, the initial condition, its numerical determination is very simple and represents a progress compared to the complete problem. Furthermore, it is easy to see that the asymptotic behaviour of *ν*_*i*_(*τ*) as *τ* → ∞ is

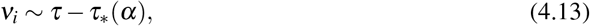

as illustrated in Fig. 3. The function *τ*_∗_ (*α*) is a universal function that can be determined numerically once and for all. A good numerical fit, with absolute error of less than 0.013 in the range 0 *< α <* 2.5, is given by

**FIG. 3.**
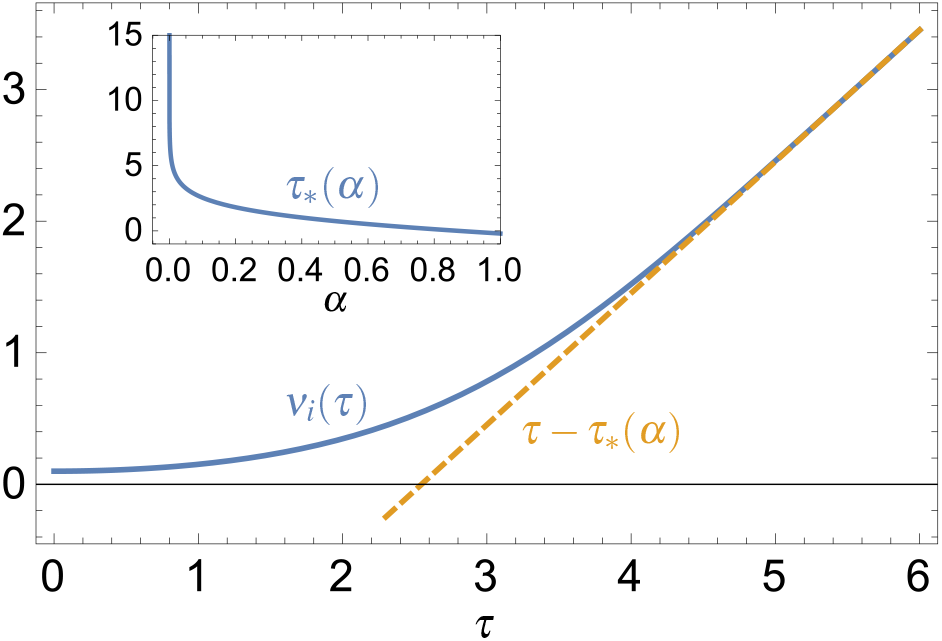
Solution of Eq. (4.10) with 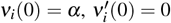, and *α* = 0.1. Dashed line, asymptotic behaviour of *ν*_*i*_(*τ*). Inset: the function *τ*_∗_(*α*).

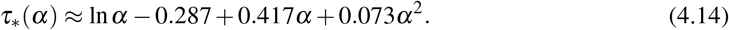

### 4.3 Matching

As *τ* → ∞, we have, combining Eqs. (4.8) and (4.13):

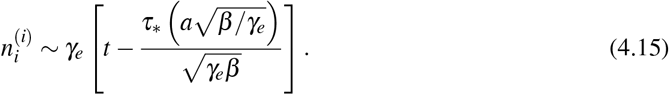

On the other hand, as *t* → *t*_*o*_, the outer solution yields

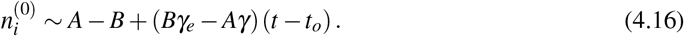

The matching conditions are, therefore,

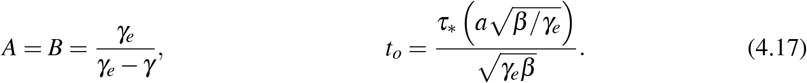

The common part between the inner and outer solutions is simply *γ*_*e*_(*t − t*_*o*_). We now have all the elements to construct a uniformly valid composite asymptotic approximation of *n*_*i*_(*t*).

For *t < t*_*o*_, only the inner solution exists because the outer solution yields negative values of *n*_*i*_:

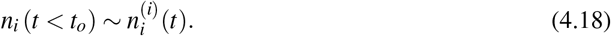

For *t > t*_*o*_, the composite solution is

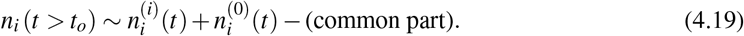

Eventually, we obtain

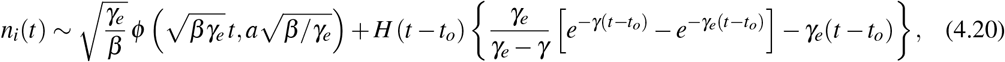

where *H*(*t*) is the Heaviside step function.

A comparison between these asymptotic approximations and the numerical solution is given in Fig. 4. Note that a much larger value of R_0_ is required to obtain a good quantitative agreement. This is because *β* and, hence, R_0_ always appear in square roots. Hence, contrarily to the SIR model, the small parameter of the problem is not 1*/*R_0_ but 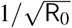. Moreover, the fast scale of the problem is not *βt* anymore but 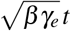. It is remarkable that the mere introduction of an intermediate variable such as the exposed population *E*(*t*) completely modifies the times scales of the dynamics, even though *γ*_*e*_ is of the same order of magnitude as *γ*.

**FIG. 4.**
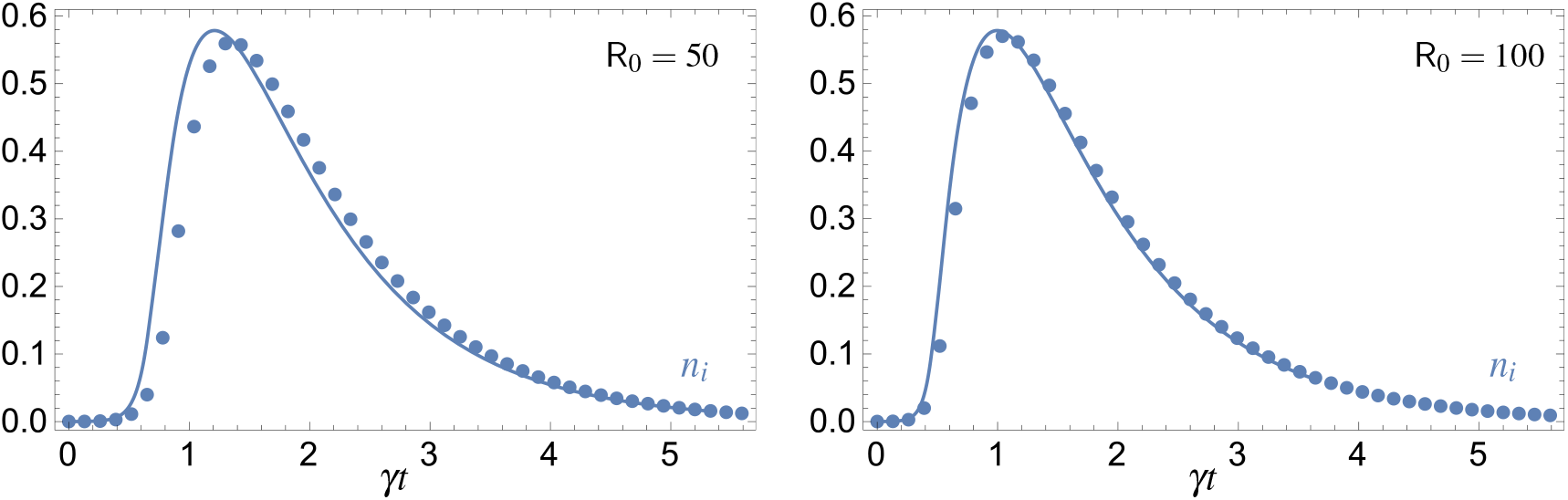
Comparison between the approximate formulas (4.20) (full lines) and the numerical solution of the SEIR equation (dotted lines) for large values of the reproduction number R_0_. *n*_*i*_(0) = *a* = 0.0001, *n*_*r*_(0) = 0, *n*_*e*_(0) = 0. The time unit is chosen so that *γ* = 1, *γ*_*e*_ = 3 and *β* = R_0_.

## 5. Conclusion

We have shown that a compact and explicit approximate solution of the SIR model can be derived in the limit of large R_0_. The interest of this solution is that it is already effective when the reproduction number is only moderately large. Given that SIR is in itself a crude approximation of reality, it makes little sense from an applied perspective to look for an approximation that is exact to within 1%. Taking the time between inflexion and maximum of the epidemic curve as a measure of accuracy, the present theory, Eq. (1.9) yields an answer with less than 10% error for R_0_ just above 4. With influenza, a reproduction number exceeding the value of 3 appears rare but possible White et al. (2009), as was the case with the 1918 influenza pandemic Mills et al. (2004). In the case of the COVID-19 pandemic estimates yield R_0_ in the range 1.5 to 6.49 Liu et al. (2020) and particularly around 3 in France Nadeau et al. (2021) and between 3.5 and 4 in Korea and in the Hubei province (China) during the first wave Choi & Ki (2020). For measles, R_0_ can be on the order of 10 or even larger Guerra et al. (2017). Hence, the large-R_0_ limit considered here is relevant, even though, of course, the assumption of a constant ratio *β/γ* does not apply to the COVID-19 pandemic. While the formulas derived in this paper appear accurate enough, it is to be noted that they can be improved. Only leading-order expressions have been used for the inner and outer solutions that apply to the ascending and receding phases of the outbreak, respectively. The as perturbations. The same can be done with 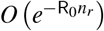terms in the equation for the outer solution. inner solution can be improved by the keeping *O* (1*/*(R_0_ − 1)) corrections in Eq. (3.4) and treating them Even within the present theory, a correction to *t*_2_ could be computed by perturbation, thus improving Eq. (1.9). In this paper, we applied the technique of matched asymptotic expansions to the SIR model.

This technique could in principle be applied to more general epidemiological models, leading to simple and reliable expressions in the limit of large reproduction numbers. We demonstrated this point with the SEIR model. Interestingly, the analysis shows that the mere introduction of the population *E* of “exposed”, not yet contagious, individuals profoundly alters the times scales of the outbreak. Also, in this second example the effective small parameter is R_0_^−1*/*2^ instead of R_0_^−1^ and so the error committed in retaining only the leading order approximation is *O* (R_0_^−1*/*2^).

## Data Availability

No data used

## Aknowledgement

G.K. is a Research Associate with the Belgian Fonds de la Recherche Scientifique (FNRS).

## A. Derivation of formulas (1.4) and (1.5)

From Eq. (2.1), d*n*_*i*_*/*d*t* vanishes when 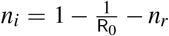. By substitution into (1.3), one immediately obtains (1.5).

Next, let us enquire when the second time derivative of *n*_*i*_ vanishes. Writing *n*_*i*_ = *n*_*i*_ [*n*_*r*_(*t*)], let us denote

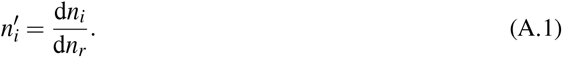

Then,

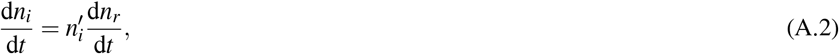

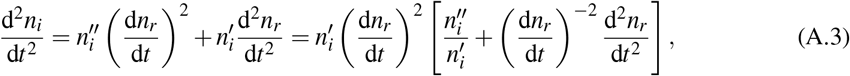

and one thus seek for the condition

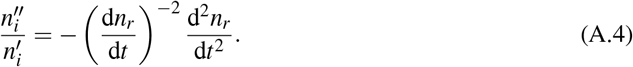

Dividing (2.1) by (2.2), one has

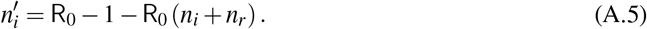

Hence, 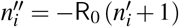 and

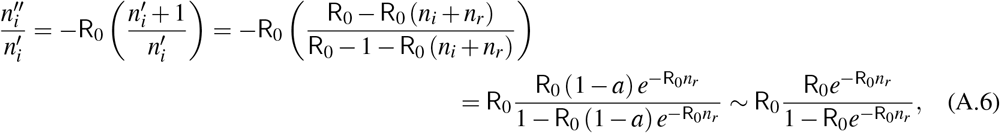

as *a* → 0. On the other hand, neglecting *a* compared to 1, (2.6) yields

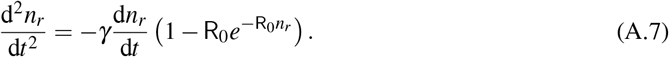

Hence,

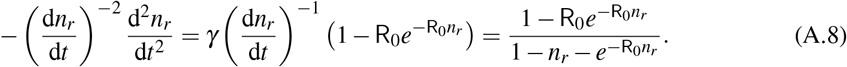

Equating (A.6) with (A.8) yields (1.4).

